# Monitoring sustainable development goal 5.2: Cross-country cross-time invariance of measures for intimate partner violence

**DOI:** 10.1101/2022.04.08.22273612

**Authors:** Kathryn M. Yount, Irina Bergenfeld, Nishat Mhamud, Cari Jo Clark, Nadine J. Kaslow, Yuk Fai Cheong

**Affiliations:** Hubert Department of Global Health and Department of Sociology, Emory University, Atlanta, GA, USA; Hubert Department of Global Health, Rollins School of Public Health, Atlanta, GA, USA; Department of Psychiatry and Behavioral Sciences, Emory University School of Medicine, Atlanta, GA, USA; Department of Psychology, Emory University, Atlanta, GA, USA

## Abstract

**Background:** The persistence and impacts of violence against women motivated Sustainable Development Goal (SDG) 5.2 to end such violence. Global psychometric assessment of cross-country, cross-time invariance of items measuring intimate partner violence (IPV) is needed to confirm their utility for comparing and monitoring national trends.

**Methods:** Analyses of seven physical-IPV items included 377,500 ever-partnered women across 20 countries (44 Demographic and Health Surveys (DHS)). Analyses of five controlling-behaviors items included 371,846 women across 19 countries (42 DHS). We performed multiple-group confirmatory factor analysis (MGCFA) to assess within-country, cross-time invariance of each item set. Pooled analyses tested cross-country, cross-time invariance using DHSs that showed configural invariance in country-level multiple-group confirmatory factor analysis (MGCFAs). Alignment optimization tested approximate invariance of each item set in the pooled sample of all datasets, and in the subset of countries showing metric invariance over at least two repeated cross-sectional surveys in country-level MGCFAs.

**Results:** In country-level MGCFAs, physical-IPV items and controlling-behaviors items functioned equivalently in repeated survey administrations in 12 and 11 countries, respectively. In MGCFA testing cross-country, cross-time invariance in pooled samples, neither item set was strictly equivalent; however, the physical-IPV items were approximately invariant. Controlling-behaviors items did not show approximate cross-country and cross-time invariance in the full sample or the sub-sample showing country-level metric invariance.

**Conclusion:** Physical-IPV items approached approximate invariance across 20 countries and were approximately invariant in 11 countries with repeated cross-sectional surveys. Controlling-behaviors items were cross-time invariant within 11 countries but did not show cross-country, cross-time approximate invariance. Currently, the physical-IPV item set is more robust for monitoring progress toward SDG5.2.1, to end IPV against women.

## Introduction

One third of women experience intimate partner violence (IPV) in their lifetime (1). IPV has a range of well-documented adverse effects on women’s mental health (2), physical health (3), and socioeconomic well-being (4), as well as effects on children (5) that perpetuate an inter-generational cycle of violence (6). The global cost of IPV against women is more than $4.4 trillion or almost 5.2% of global gross domestic product (7).

The high prevalence, adverse effects, and persistence of IPV have motivated many calls to end violence against women particularly in their intimate relationships. A landmark commitment to end IPV against women was embodied in Sustainable Development Goal Target 5.2, which calls on national governments to “eliminate all forms of violence against all women and girls in public and private spheres, including trafficking and sexual and other types of exploitation” (8). Indicator 5.2.1 is defined to measure the “proportion of ever-partnered women and girls aged 15 years and older subjected to physical, sexual or psychological violence by a current or former intimate partner in the previous 12 months, by form of violence and by age” (1).

The global commitment to monitor this indicator has generated a surge in research to understand the measurement properties of questionnaire modules assessing the major dimensions of IPV against women. Studies using survey data from 28 European Union (EU) countries have established the strict measurement invariance of measures for psychological (9), sexual (10), and physical (10) IPV. These studies relied on measures specific to the EU survey, which include more items for physical IPV (10 items), sexual IPV (4 items), controlling behaviors (8 items) and other psychological IPV (5 items) than used in other cross-national surveys that collect similar data. In low- and middle-income countries (LMICs), commonly used modules to measure IPV come from the World Health Organization (WHO) Multi-Country Study of Women’s Health and Domestic Violence (11) and the Demographic and Health Survey (DHS) Domestic Violence Module (DMV) (12). The DHS DMV, which has aligned with the WHO IPV module, includes items to measure physical IPV (7 items) and controlling behaviors (5 items) in 89 DHS spanning 58 countries from 2005 to 2020. A recent analysis tested the cross-national invariance of the physical-IPV items and controlling-behaviors items for 36 countries for the period 2012-2018. Findings demonstrated approximate invariance of both item sets (13).

An important step to confirm the utility of these items for monitoring SDG 5.2.1 is to assess their *cross-*national and cross-time-invariance. Such an analysis would ascertain the measurement properties of these items both across countries and across repeated national surveys conducted with some periodicity. Evidence of the joint cross-national and cross-time invariance of these items would provide even stronger evidence for our capacity to monitor SDG 5.2.1. To date, the cross-national and cross-time invariance of these items is unknown, and the analysis presented here is designed to fill that critical gap. Our primary objective was to assess the cross-national and cross-time invariance of the seven DHS physical-IPV items, and separately, the five DHS controlling-behaviors items for the 20 and 19 countries, respectively, that had administered at least two repeated cross-sectional DHS approximately five years apart.

## Methods

### Sample and data on IPV

The DHS is a U.S. Agency for International Development (USAID)-funded program operating across more than 90 LMICs that collects data on population and health, including IPV among ever-partnered women. Per DHS protocols, between 15% and 100% of sampled households are administered a DVM, for which one woman 15-49 years in the household is randomly selected and interviewed (Table 1). To ensure similarity in the number and wording of items across administrations of the DHS, we restricted our sample to DVM versions V through VII, administered between 2005 and 2019. Within this frame, final samples included ever-partnered women 15-49 years from 19 countries and 18-49 years from one country in which at least two DHS were administered at intervals of 1 to 12 years with the same seven physical-IPV items (44 surveys total). For analyses of the controlling-behaviors items, two Rwanda surveys in the above sample were excluded; one survey did not administer the controlling behaviors items, bringing the number of countries to 19 and the number of surveys to 42.

**Table 1.**
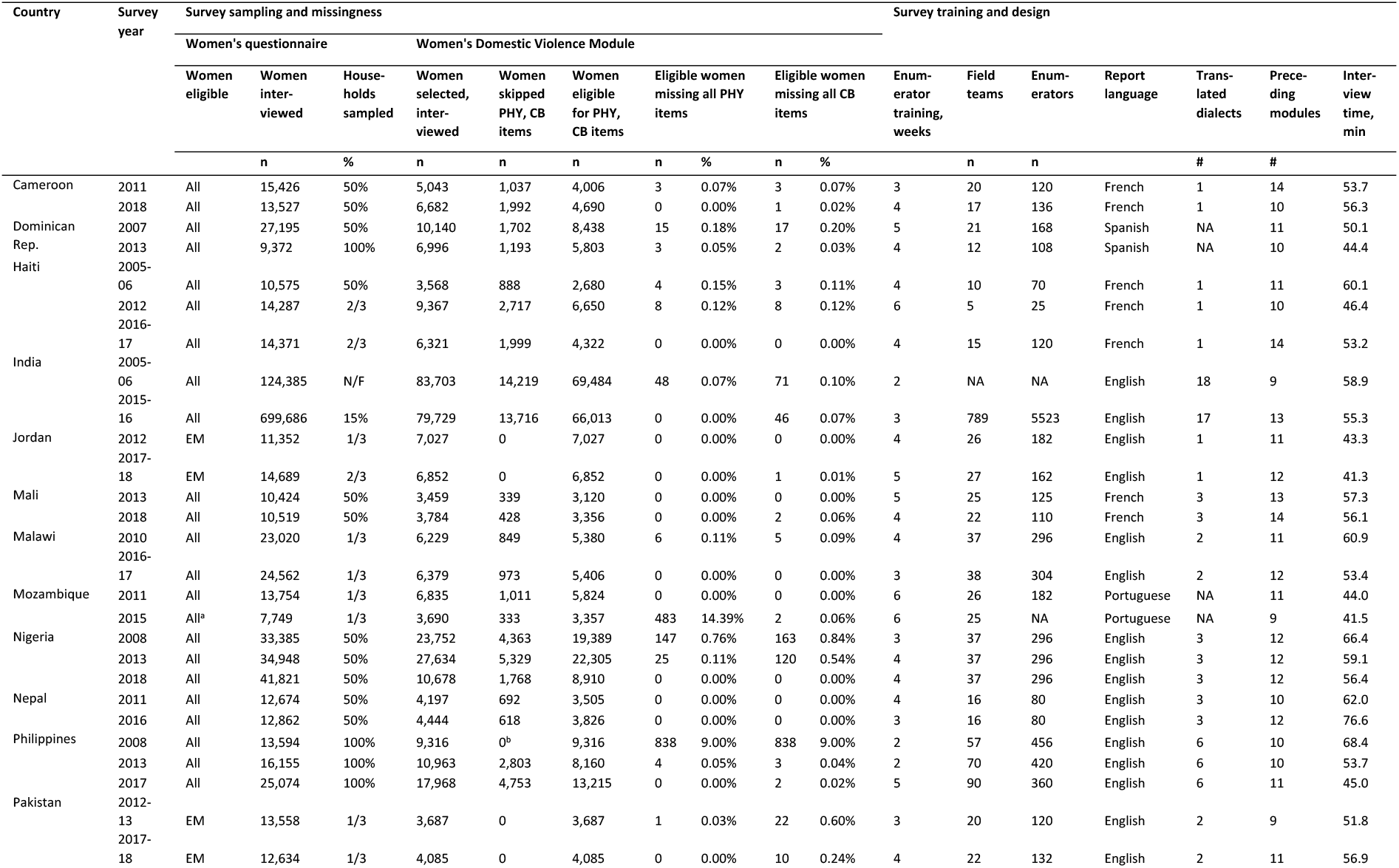
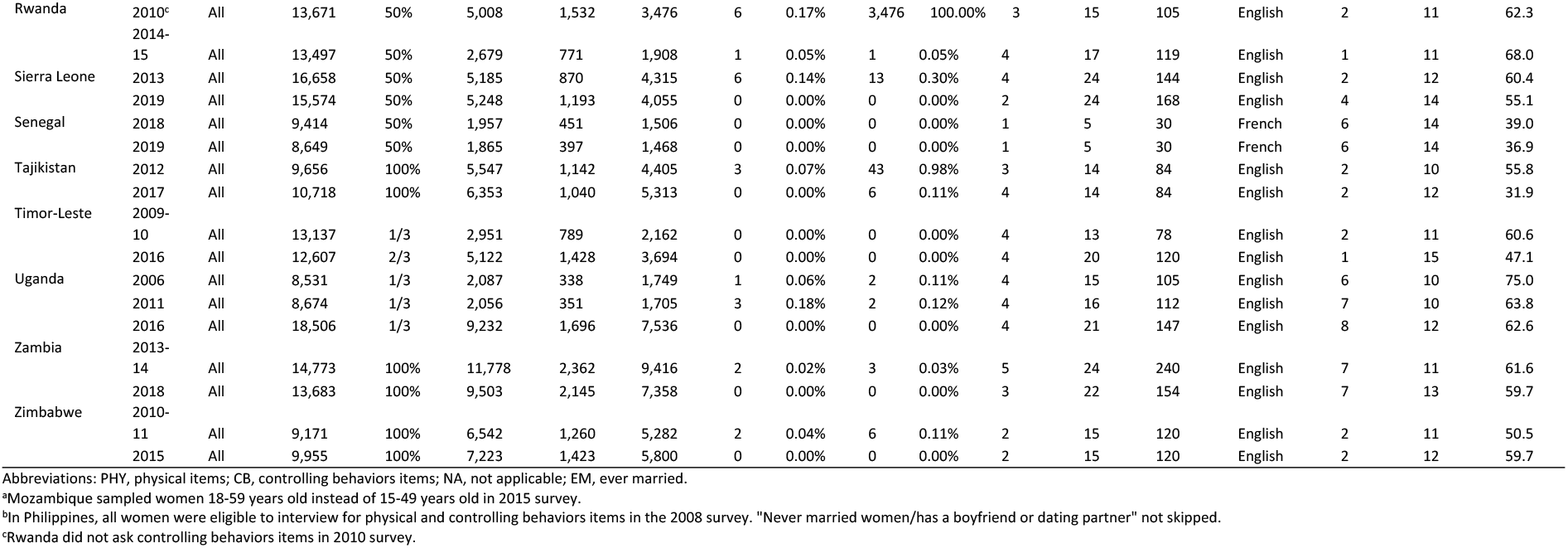
Countries, Demographic Health Survey samples, and intimate partner violence items included in time-invariance analysis.

The total sample included 380,012 women who were selected and administered the DVM and were not skipped out of the IPV items due to never-partnered status. Of these, 2,512 were missing data on all physical-IPV items, bringing the final analytic sample to 377,500 ever-partnered women across 20 countries and 44 DHS in analyses of the physical IPV items. The final analytic sample for controlling-behaviors items was 374,628 women across 19 countries and 42 DHS. Removal of individuals with missing and “don’t know” responses for all controlling behaviors items brought the analytical sample for controlling behaviors to 371,846. All DHS samples were downloaded with written permission from the DHS program.

For items on physical, sexual, and psychological IPV, participants across all DHS were asked whether their husband or partner had ever done each act (see Supplemental Table S1 for item wordings). Participants who responded yes were asked whether their husband or partner had done the act often, sometimes, or never within the past 12 months. For controlling behaviors, participants were asked whether their husband or partner did or did not do each of the behaviors without reference to a time frame. We elected to use the lifetime rather than the past-12-month timeframe for responses to the physical-IPV items for greater comparability across the two item sets. In a subset of countries, the DHS also included a maximum of three sexual-IPV items and a maximum of three psychological-IPV items (Supplemental Table S1). We did not use these item sets in our final analyses due to their questionable content validity relative to uniform definitions of these constructs (14, 15) and the small number of included items (16).

### Analytic strategy

In step 1 we tested the measurement invariance of the set of seven physical-IPV items, and separately, the five controlling-behaviors items, over repeated cross-sectional surveys within each country. We performed multiple group confirmatory factor analysis (MGCFA) using weighted least squares estimation, comparing the fit of configural models, in which all loadings and thresholds were estimated freely across repeated cross-sectional surveys, and scalar models, in which all loadings and thresholds were constrained to be equal across repeated cross-sectional surveys. For countries with three repeated cross-sectional surveys, we performed invariance testing across each combination of two surveys. We used DHS-generated probability weights and cluster variables in all models to account for selection probabilities and clustering. We used several indices to assess the fit of configural models: chi-square (χ^2^), Root Mean Square Error of Approximation (RMSEA, adequate fit ≤0.08, good fit ≤0.05), and Comparative Fit Index and Tucker-Lewis Index (CFI, TLI, ≥0.95) (17). We used the χ^2^ difference test to assess invariance over repeated cross-sectional surveys (18, 19).

In step 2, we conducted a pooled analysis of all DHSs that showed configural invariance in each individual-country MGCFA. In step 3, we used MGCFA with maximum likelihood (ML) estimation to assess metric invariance across repeated cross-sectional surveys within each country and in a pooled analysis across all DHSs. When pooled analyses showed a lack of evidence for metric invariance, we used the alignment optimization (AO) approach in step 4 to perform an approximate invariance test in the pooled sample of all datasets. This approach relaxes some assumptions of MGCFA by allowing estimated country-specific model parameters to vary from the estimated model parameters in the pooled dataset following a normal distribution. The criterion for approximate invariance is evidence that ≤25% of model parameters (loadings and thresholds) are non-invariant. In step 5, where approximate invariance was not supported in the pooled sample of all datasets, we restricted the sample to countries that showed metric invariance over repeated cross-sectional surveys in individual-country analyses. We then reran alignment optimization using this subset of surveys and countries. We used STATA 17 (20) for data cleaning and management. All measurement invariance testing was performed in MPlus 8 (21).

## Results

### Characteristics of included surveys

Survey characteristics, including logistics and design, are summarized across the full sample of 44 surveys (Table 1). The duration of enumerator training varied across surveys from between one to six weeks, with most surveys (43%) conducting training in four weeks. Across all surveys, data collection was conducted by an average of 42 field teams. The total number of survey field teams ranged from five in Senegal to 789 in India. Most surveys (91%) were translated into at least one local dialect. India had the most translations, into 17-18 dialects. All surveys included three sensitive modules on HIV, contraception, and sexual activity that preceded the DVM. The DVM typically was the last module in the women’s questionnaire, with at least nine modules preceding it. Nearly half of the surveys (21 of 44) reported a mean duration of the women’s interview of 45 to 60 minutes; however, the interview duration ranged from 31.9 minutes to 76.6 minutes.

Surveys were administered between 2005 and 2019; 16% were administered in 2018. Within countries, the average number of years between repeated survey administrations was five. To create the DVM sample, all surveys selected between 15% and 100% of households interviewed in the main DHS, and then sampled one woman per household for the DVM. A plurality of surveys (39%) sampled 50% of interviewed households to create the household sample for the DVM. On average, 10,520 women across surveys were selected and interviewed for the DVM; however, sample sizes for the DVM ranged from 1,865 women in the 2019 Senegal DHS to 83,703 women in the 2005-06 India DHS. In most surveys (91%), both ever-married and never-married women were eligible for the DVM. Two surveys in Jordan and two surveys in Pakistan interviewed only ever-married women for the DVM. Among the surveys that interviewed all women for the DVM, only ever-married women and women who ever lived with a man were eligible for the physical IPV and the controlling behaviors items. However, in the 2008 Philippines DHS, women who have (had) a boyfriend or dating partner previously or at the time of the survey were eligible for the physical-IPV and controlling-behaviors items. All surveys interviewed women ages 15 to 49 years for the DVM, except the 2015 Mozambique DHS, which included women 18 to 59 years.

### Country-specific and pooled time invariance of physical IPV items and controlling behaviors items

Of the 20 countries included in the measurement-invariance testing of the seven physical-IPV items, all showed good fit for the individual-country configural model across at least two DHS administrations (Table 2). According to changes-in-fit-statistics criteria (ΔCFI, ΔTLI), individual-country models showed scalar invariance over time; however, according to the χ2 difference test between scalar and configural models, individual-country models for only five countries showed scalar invariance over time. In metric invariance testing using maximum likelihood estimation, 12 countries had a non-significant likelihood ratio test across repeated DHS administrations in individual-country analyses, suggesting metric invariance (Table 3).

**Table 2.**
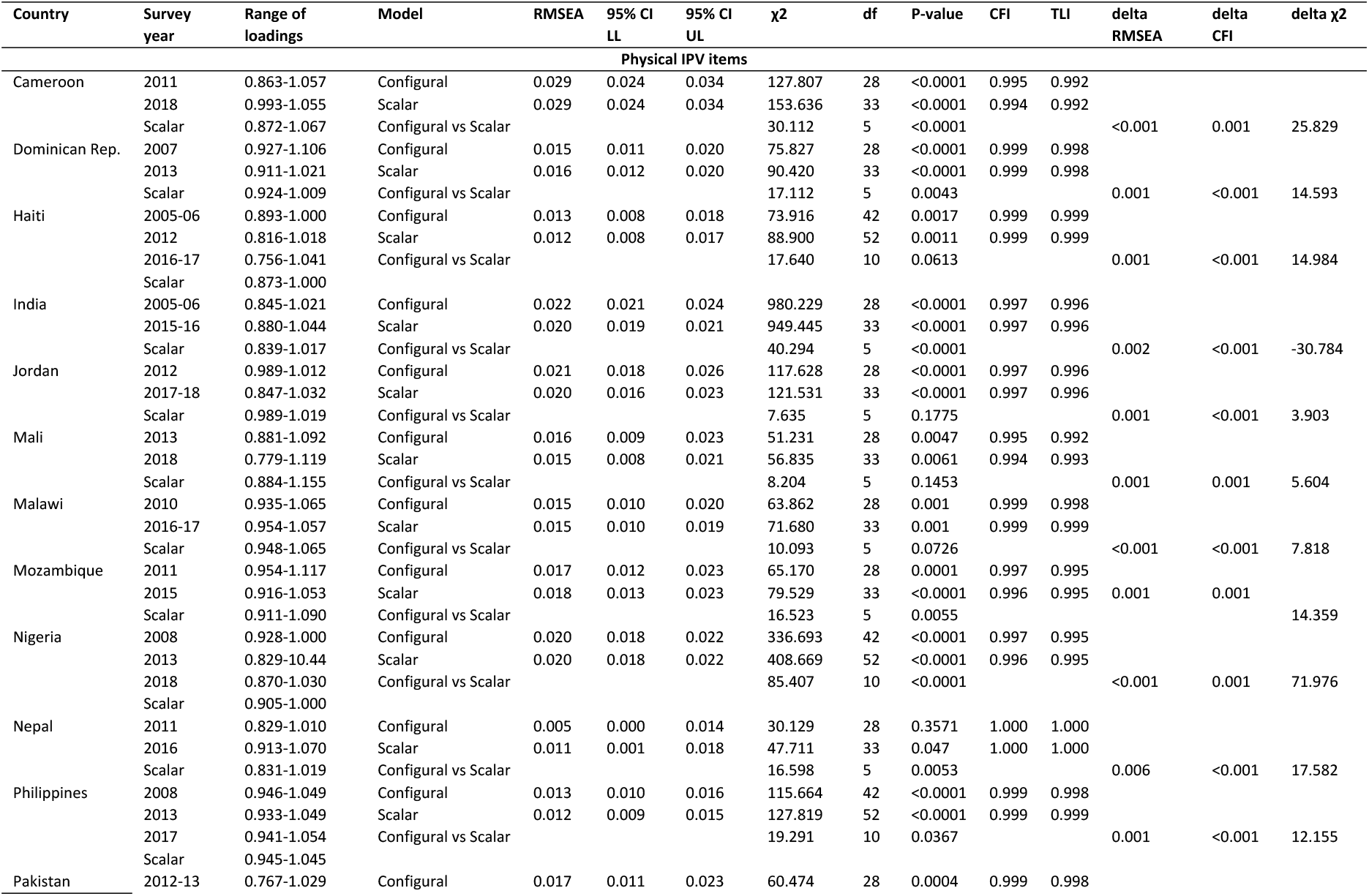
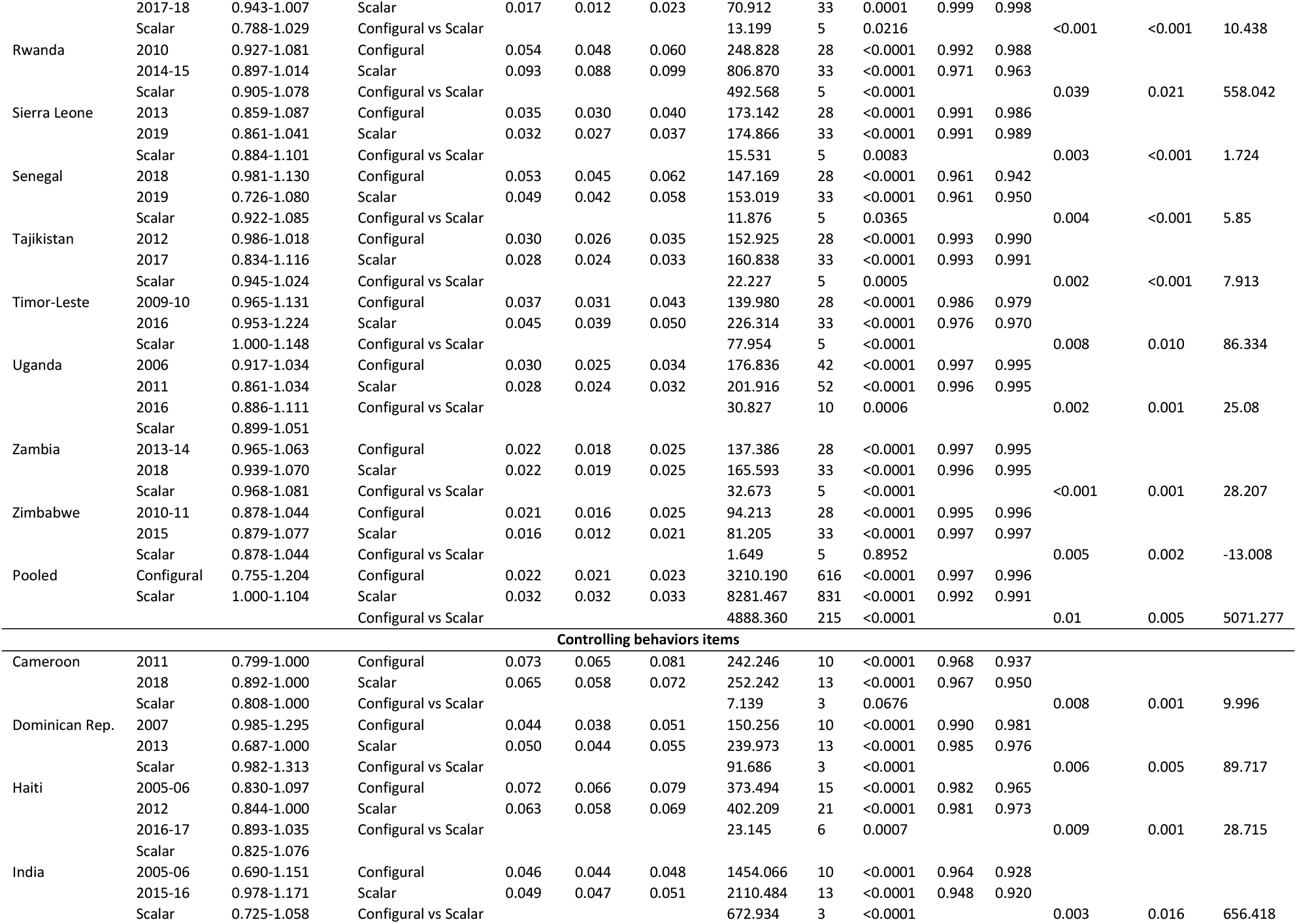
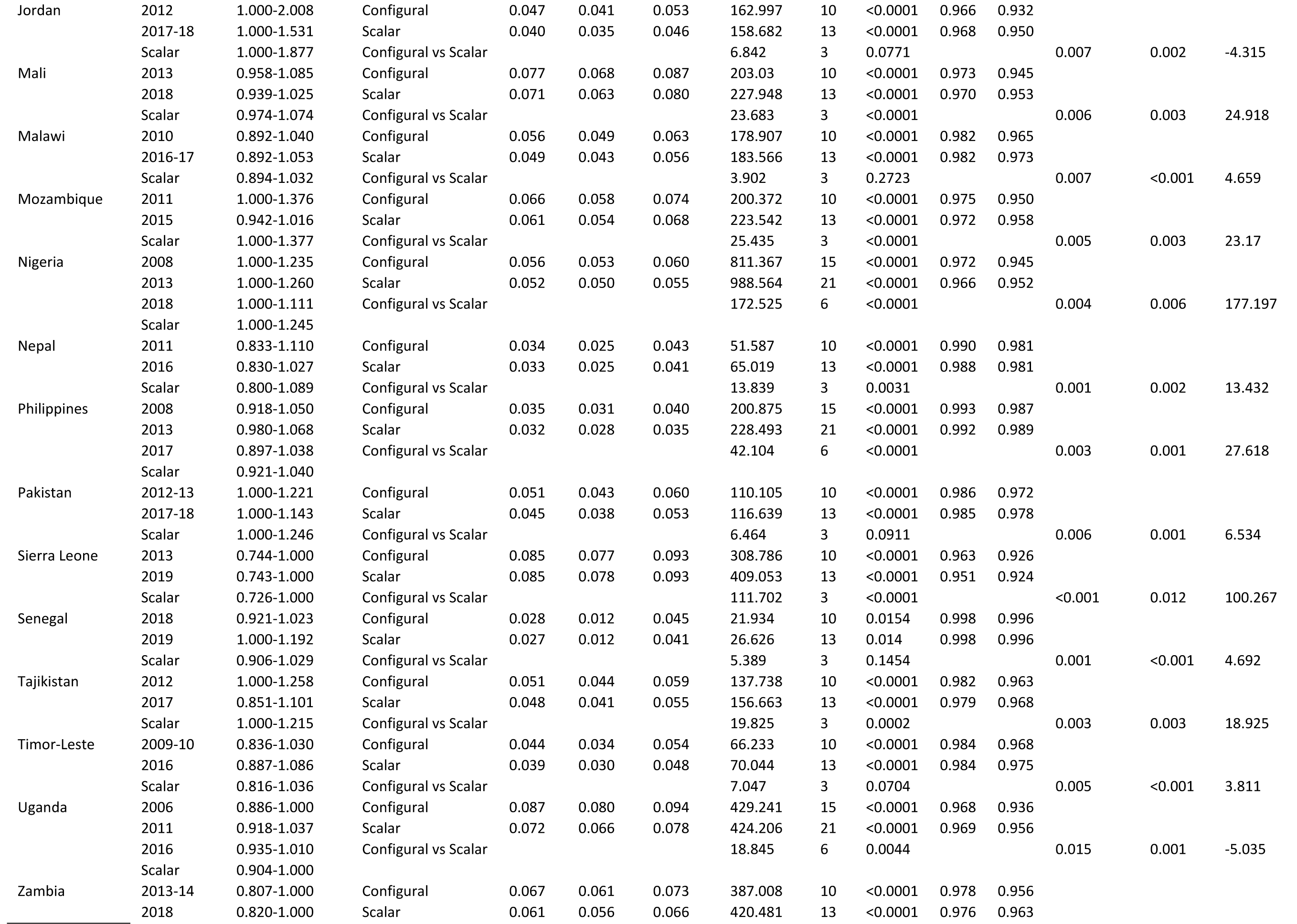
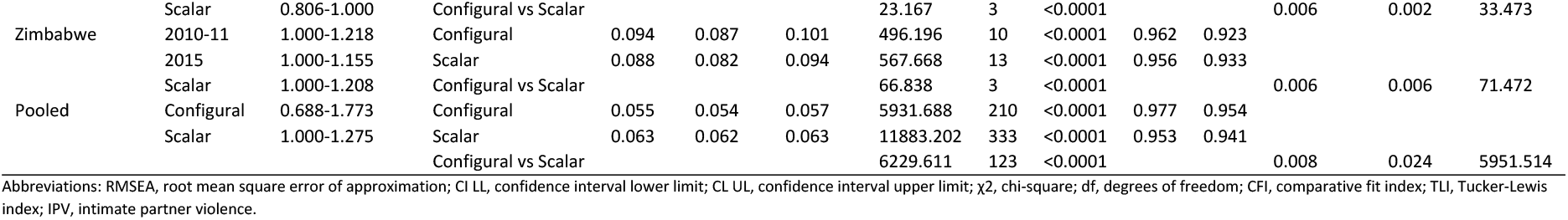
Scalar invariance testing for Demographic Health Survey physical intimate partner violence items (n=20 countries) and controlling behaviors Items (n=19 countries).

**Table 3.**
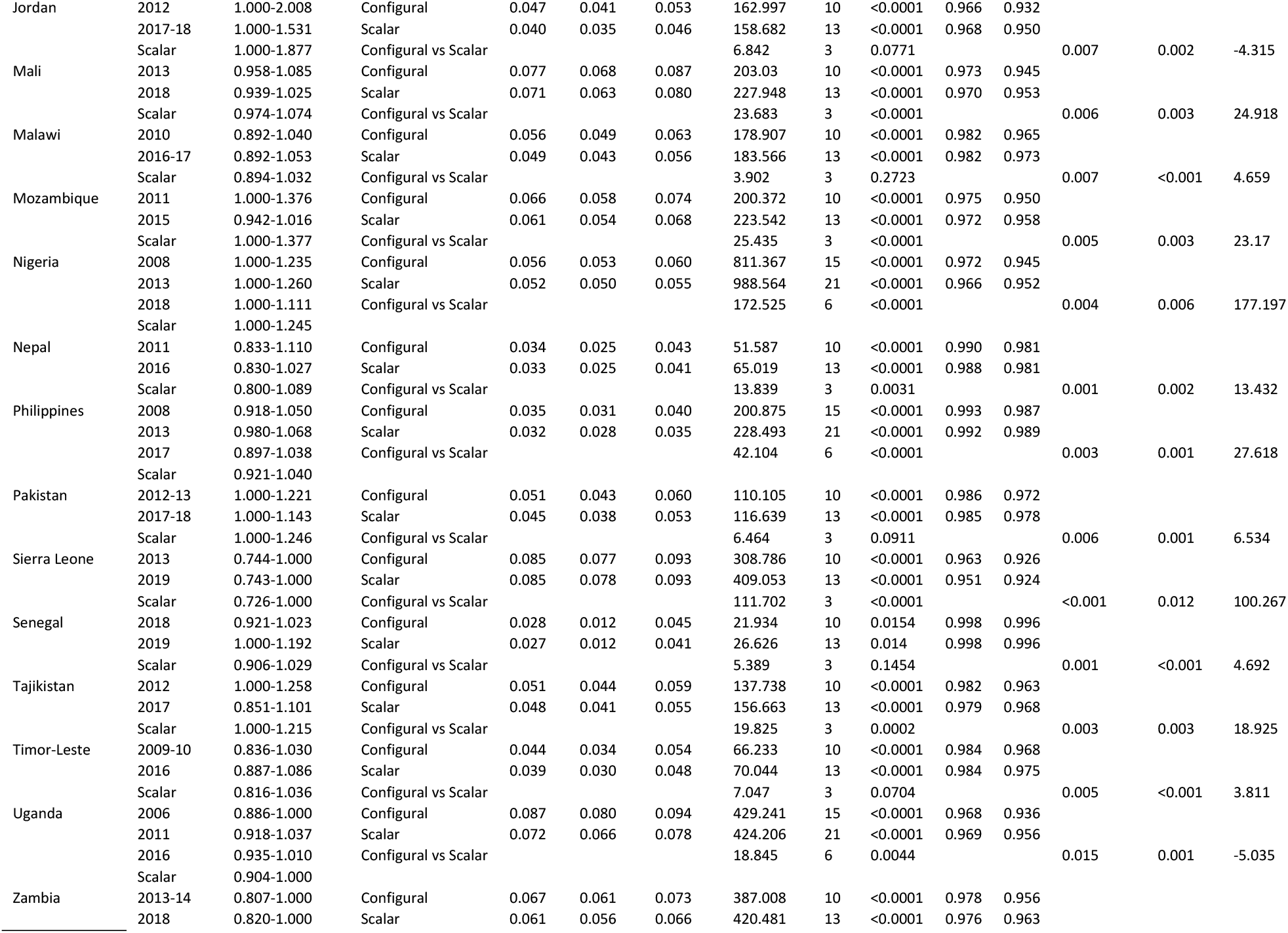
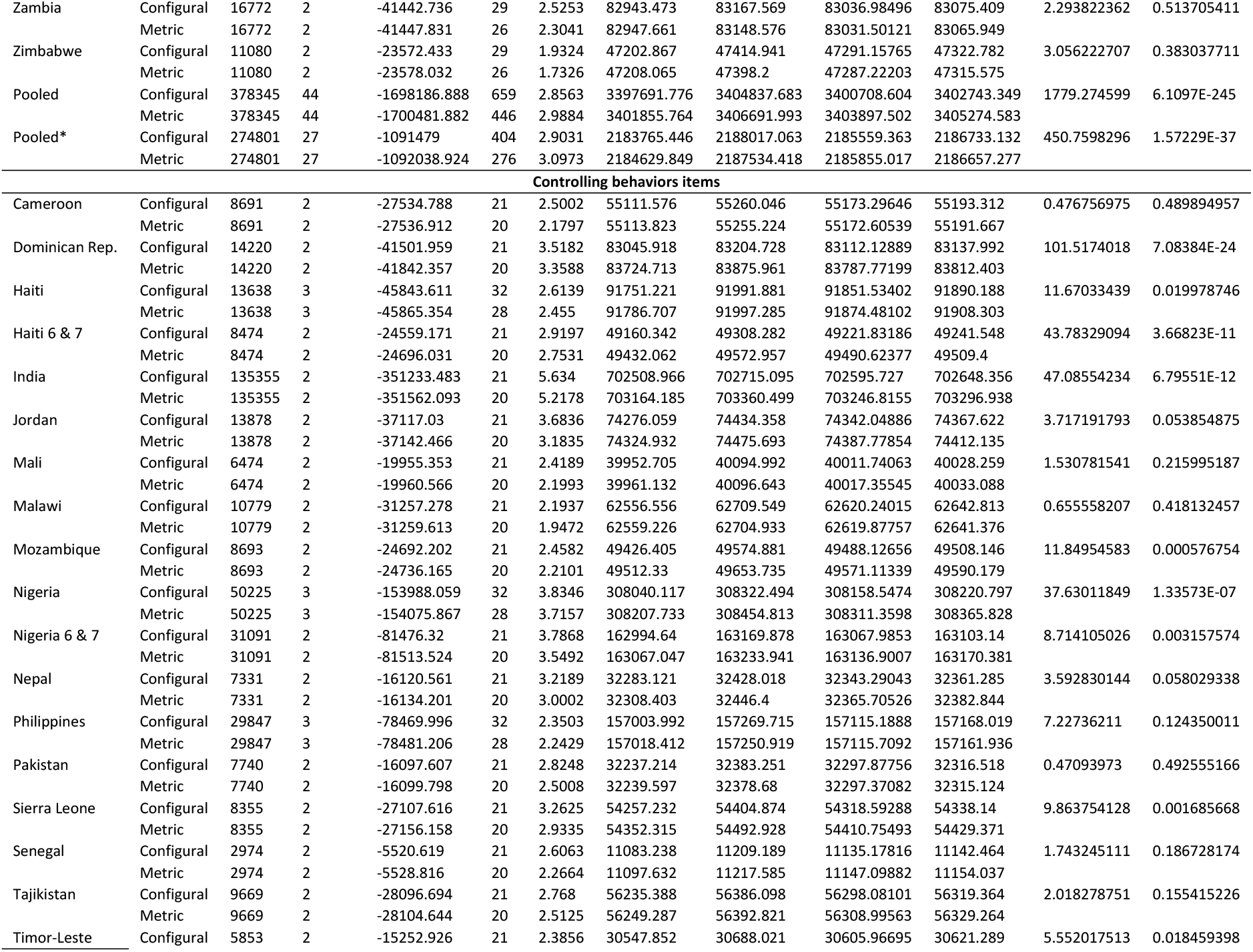
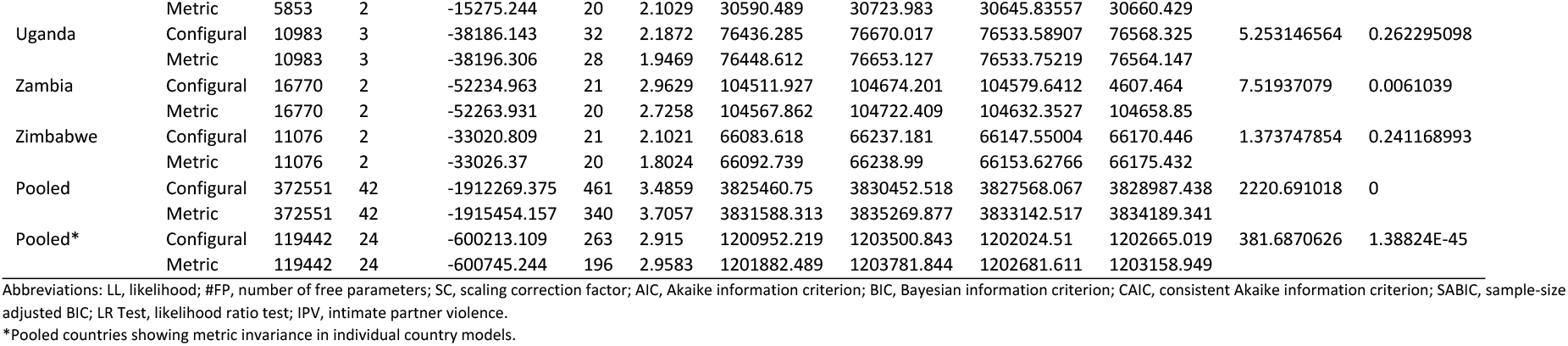
Metric invariance testing for Demographic Health Survey physical intimate partner violence items (n=20 countries) and controlling behaviors items (n=19 countries).

In a pooled analysis of all 20 countries, while configural invariance was evident, neither metric (Table 3) nor scalar (Table 2) invariance was achieved according to difference testing. Changes in fit statistics, however, did not provide evidence of non-invariance in the scalar models. When the pooled sample was restricted to the 27 DHSs (from 12 countries) that showed evidence of metric invariance across repeated DHS administrations in within-country analyses, metric invariance still was not evident (Table 3).

In analyses of the five controlling-behaviors items, all 19 individual-country analyses showed good fit of the configural model (Table 2). Six countries showed evidence of scalar invariance over time according to χ2 difference testing, with 11 showing evidence of metric invariance according to the likelihood ratio test in maximum likelihood models (Table 3). For the pooled sample of 19 countries, neither metric nor scalar invariance was suggested by the likelihood ratio test or the χ2 difference test, respectively. Metric invariance was not evident in the 11-country pooled sample for which repeated cross-sectional DHSs showed metric invariance over time in individual-country analyses. Neither the individual-country nor pooled analyses showed evidence of non-invariance according to changes in fit statistics in weighted least squares models.

### Tests of approximate invariance of physical-IPV items and controlling-behaviors items

Table 4 presents the AO-based results, in which we assessed approximate measurement invariance separately for the seven physical-IPV items (Panel 1) and the five controlling-behaviors items (Panel 2). For physical IPV, 118 (or 38% of) estimated thresholds, 44 (or 14% of) estimated loadings, and 26% of all parameter estimates were measurement non-invariant (Supplemental Table S2). For controlling behaviors, 132 (or 61% of) estimated thresholds, 78 (or 36% of) estimated loadings, and 49% of all parameter estimates were measurement non-invariant. A guideline of 25% or fewer total non-invariant parameter estimates is recommended for trustworthy latent mean estimates and their comparison across groups. The results suggested that neither item set exhibited approximate measurement invariance across the 20 countries and repeated DHS administrations. Among the seven physical-IPV items, the item ‘slap’ had a low degree of threshold and loading invariance, as shown by its low R^2^ (Table 4).

**Table 4.** Thresholds, loadings, and R^2^ values from alignment optimization analysis of physical intimate partner violence items and controlling behaviors items using the full pooled sample of Demographic and Health Surveys.

| **Panel A. Results from alignment optimization analysis for physical Items, n=378,345 across Demographic Health Surveys in 20 countries, 2006-2019** |  |  |  |  |
| --- | --- | --- | --- | --- |
| Items | Thresholds |  | Loadings |  |
| | Weighted average value across invariant groups | $R^2$ | Weighted average value across invariant groups | $R^2$ |
| Push you, shake you, or throw something at you? | 2.25 | 0.589 | 2.946 | 0.346 |
| Slap you? | 0.09 | 0.011 | 3.511 | 0.00 |
| Punch with his fist or with something that could hurt you? | 3.362 | 0.528 | 3.217 | 0.618 |
| Kick you, drag you, or beat you up? | 4.473 | 0.662 | 3.11 | 0.635 |
| Try to choke you or burn you on purpose? | 5.916 | 0.546 | 2.599 | 0.509 |
| Threaten to attack you with a knife, gun or other weapon? | 5.85 | 0.459 | 2.602 | 0.511 |
| Twist your arm or pull your hair? | 3.585 | 0.756 | 2.919 | 0.546 |
| # (%) of threshold non-invariant parameters = 118 (38) |  |  |  |  |
| # (%) of loading non-invariant parameters = 44 (14) |  |  |  |  |
| # (%) of total non-invariant parameters = 162 (26) |  |  |  |  |
| **Panel B. Results from alignment optimization analysis for controlling behaviors Items, n=372, 692 across Demographic Health Surveys in 19 countries, 2006-2019** |  |  |  |  |
| Items | Thresholds |  | Loadings |  |
| | Weighted average value across invariant groups | $R^2$ | Weighted average value across invariant groups | $R^2$ |
| Is jealous or angry if she talks to other men? | -1.653 | 0.759 | 2.457 | 0.179 |
| Frequently accuses her of being unfaithful? | 1.116 | 0.846 | 2.359 | 0.309 |
| Does not permit her to meet her female friends? | 1.266 | 0.706 | 2.835 | 0.406 |
| Tries to limit her contact with her family? | 3.422 | 0.627 | 2.984 | 0.426 |
| Insists on knowing where she is at all times? | -0.216 | 0.824 | 1.921 | 0.594 |
| # (%) of threshold non-invariant parameters = 132 (61) |  |  |  |  |
| # (%) of loading non-invariant parameters = 78 (36) |  |  |  |  |
| # (%) of total non-invariant parameters = 210 (49) |  |  |  |  |

Table 5 presents the AO-based results in which we assessed approximate measurement invariance separately for the physical-IPV items and the controlling-behaviors items for a subset of countries that displayed metric invariance across at least two administrations of the DHS (Table 5). For physical IPV, 61 (or 36% of) estimated thresholds, 15 (or 9% of) estimated loadings, and 20% of all parameter estimates were measurement non-invariant. For controlling behaviors, 47 (or 39% of) estimated thresholds, 21 (or 17.5% of) estimated loadings, and 28% of all parameter estimates were measurement non-invariant. Thus, the results suggested that DHS physical-IPV items but not the controlling-behaviors items exhibited approximate measurement invariance across countries and repeated administrations showing within-country metric invariance and allowed acceptable alignment performance. Additionally, the R^2^ values showed that all seven physical-IPV items had a reasonable degree of threshold and loading invariance (Table 5).

**Table 5.**
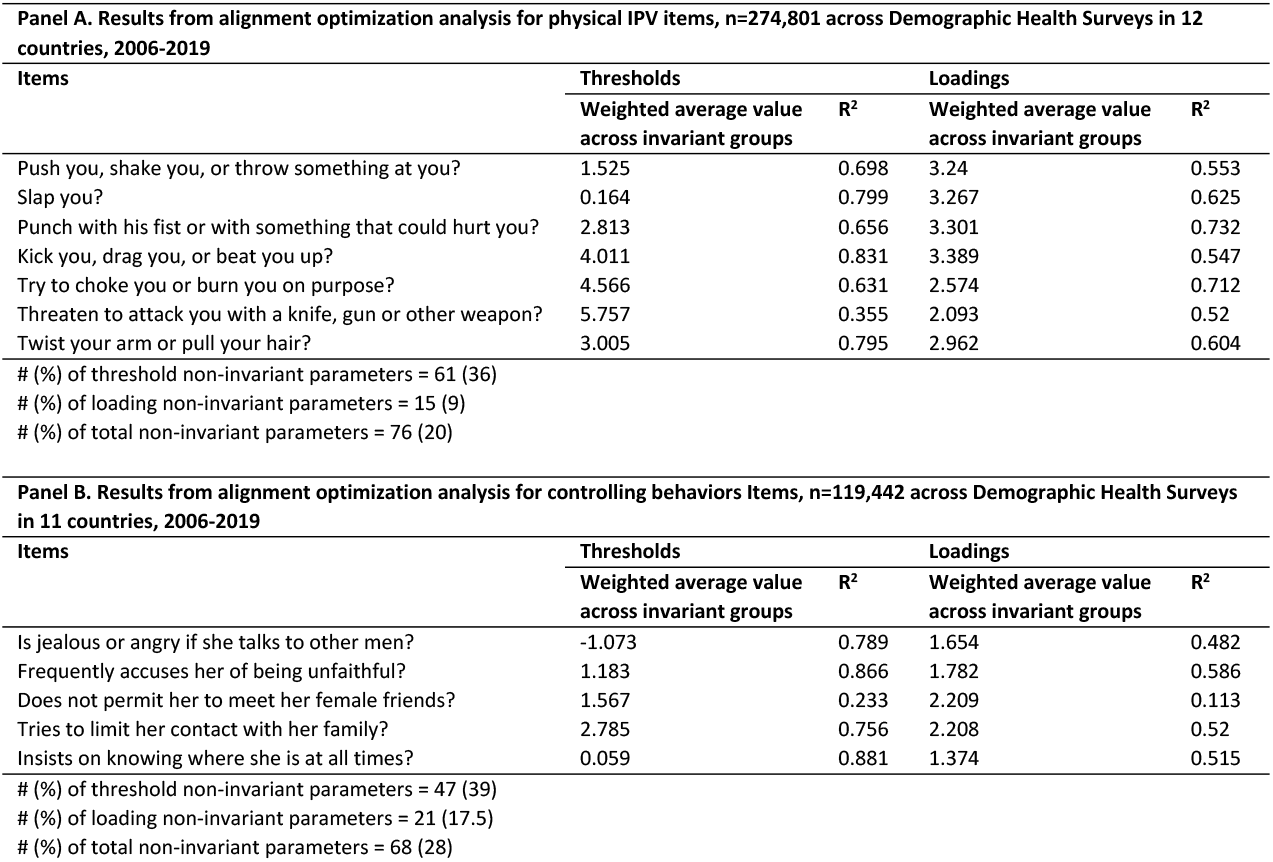
Thresholds, loadings, and R^2^ values from alignment optimization analysis of physical IPV items and controlling behaviors items using the subsetted pooled sample of Demographic and Health Surveys.

| Panel A. Results from alignment optimization analysis for physical IPV items, n=274,801 across Demographic Health Surveys in 12 countries, 2006-2019 |  |  |  |  |
| --- | --- | --- | --- | --- |
| Items | Thresholds |  | Loadings |  |
| | Weighted average value across invariant groups | $R^2$ | Weighted average value across invariant groups | $R^2$ |
| Push you, shake you, or throw something at you? | 1.525 | 0.698 | 3.24 | 0.553 |
| Slap you? | 0.164 | 0.799 | 3.267 | 0.625 |
| Punch with his fist or with something that could hurt you? | 2.813 | 0.656 | 3.301 | 0.732 |
| Kick you, drag you, or beat you up? | 4.011 | 0.831 | 3.389 | 0.547 |
| Try to choke you or burn you on purpose? | 4.566 | 0.631 | 2.574 | 0.712 |
| Threaten to attack you with a knife, gun or other weapon? | 5.757 | 0.355 | 2.093 | 0.52 |
| Twist your arm or pull your hair? | 3.005 | 0.795 | 2.962 | 0.604 |
| # (%) of threshold non-invariant parameters = 61 (36) |  |  |  |  |
| # (%) of loading non-invariant parameters = 15 (9) |  |  |  |  |
| # (%) of total non-invariant parameters = 76 (20) |  |  |  |  |
| Panel B. Results from alignment optimization analysis for controlling behaviors Items, n=119,442 across Demographic Health Surveys in 11 countries, 2006-2019 |  |  |  |  |
| Items | Thresholds |  | Loadings |  |
| | Weighted average value across invariant groups | $R^2$ | Weighted average value across invariant groups | $R^2$ |
| Is jealous or angry if she talks to other men? | -1.073 | 0.789 | 1.654 | 0.482 |
| Frequently accuses her of being unfaithful? | 1.183 | 0.866 | 1.782 | 0.586 |
| Does not permit her to meet her female friends? | 1.567 | 0.233 | 2.209 | 0.113 |
| Tries to limit her contact with her family? | 2.785 | 0.756 | 2.208 | 0.52 |
| Insists on knowing where she is at all times? | 0.059 | 0.881 | 1.374 | 0.515 |
| # (%) of threshold non-invariant parameters = 47 (39) |  |  |  |  |
| # (%) of loading non-invariant parameters = 21 (17.5) |  |  |  |  |
| # (%) of total non-invariant parameters = 68 (28) |  |  |  |  |

## Discussion

### Summary of findings

Testing of within-country cross-time measurement invariance, relevant for national efforts to monitor IPV trends using the DHS DVM, revealed that the seven physical-IPV items and the five controlling-behaviors items functioned equivalently in repeated survey administrations within a subset of LMIC countries. In the second stage, we examined cross-country and cross-time invariance in pooled samples including multiple countries with two or more survey administrations each. While these two item sets were not strictly equivalent in these samples, the physical-IPV item set exhibited approximate invariance over time and across countries in a restricted sample of countries exhibiting within-country, cross-time metric invariance of the item set. The five controlling-behaviors items did not meet the recommended threshold for non-invariant parameters to infer approximate invariance across time and across countries. A prior analysis found evidence of approximate invariance for physical-IPV and controlling-behaviors item sets across 36 DHS administered in 36 countries during 2012-2018 (13, 22). The present analysis corroborates that the physical-IPV items function comparably across time and across very diverse national contexts and highlights the cross-time invariance of the controlling-behaviors items within selected countries. Evidence of greater threshold than loading non-invariance, especially for controlling-behaviors items, suggests greater comparability in item interpretation across contexts and time, but less comparability in the likelihood of endorsing items (responding yes to acts of IPV) across contexts and time.

### Limitations and strengths

Findings should be interpreted considering the study’s limitations and strengths. The study assessed measurement properties of item sets in the DHS; therefore, findings cannot be extended to item sets that measure other forms of IPV nor to item sets that are used in other, non-DHS IPV survey modules. However, the DHS are widely administered across LMICs and represent approximately half the data being reported to monitor progress toward SDG5.2.1. It is the single largest contributor to SDG5.2.1 monitoring and has IPV items like those used in WHO surveys, making it possibly the most important source for rigorous psychometric testing. Findings reported here may represent a best-case scenario. The DHS program provides technical support for survey administration, which, while not entirely uniform across countries or time periods (Table 1), does provide a level of consistency in administration that does not exist across the wide variety of survey formats and forms of administration that represent the data pool available for SDG5.2.1 monitoring. This level of consistency bolsters its use for research, but potentially limits study findings to the item sets tested using similarly consistent methods of administration. Finally, in pooled analyses, we were unable to account for possible auto-correlation across national surveys within countries. Despite their limitations, the findings are based on 44 DHS conducted in 20 diverse countries spanning four regions (Africa, Asia, Latin America, and the Middle East) and 15 years (2005-2019).

### Implications for research and policy

These findings suggest the seven DHS physical-IPV items are promising for comparing and monitoring national trends in IPV toward achieving SDG5.2.1, to eliminate IPV against women. The low R^2^ for the item ‘slap’ in AO analysis suggests the potential benefit of focused cognitive testing of this item across diverse contexts to improve its measurement properties, and thereby, the item set as a whole. The five DHS controlling-behaviors items, in their present formulation, show promise in some countries for monitoring within-country trends in this form of IPV; however, their lack of approximate invariance in full and restricted pooled samples of countries with repeated DHS administrations caution against their use to compare and to monitor national trends in this form of IPV toward achieving SDG5.2.1. Cognitive testing of these items, and psychometric testing of a revised controlling-behaviors item set in diverse, multi-country samples of women, may improve their measurement properties and their utility for monitoring SDG5.2.1 cross-nationally and over time.

Improved global measures of controlling behaviors also will improve our estimates of the impacts of these forms of IPV on the health of victims and their children worldwide, providing insights into strategies for prevention and response. These advances are critical, given that controlling behaviors in an intimate partnership often indicate more severe forms of IPV. Improved measurement of controlling behaviors is motivated further by changes in some criminal codes to include ‘controlling or coercive behaviors’ as prosecutable offenses (23). Thus, promoting standard, contextually informed, definitions of controlling behaviors and enhancing the measurement properties of controlling-behaviors items will strengthen the capacity for cross-national monitoring of trends, and may stimulate changes to other national criminal codes to include controlling behaviors as a prosecutable offense. Such changes would provide new legal norms about the nature and scope of IPV and new mechanisms to deter controlling behaviors (24). Such changes also offer the potential to move away from a narrow focus on physical injury towards emotional and psychological trauma in criminal cases of IPV (23), expanding response options for victims of these forms of IPV.

This analysis did not assess the cross-country and cross-time measurement properties of DHS item sets measuring psychological IPV (typically 3 items) and sexual IPV (typically 2-3 items). Presently, these DHS item sets align only narrowly with uniform definitions of these forms of IPV (15, 25), suggesting a notable lack of content validity. Still, there may be practical benefit in future analyses to assess the psychometric properties of these limited item sets to establish an evidence-base regarding the extent of their cross-country and cross-time measurement invariance. The current lack of content validity of these item sets, however, has important practical implications for interpreting trends in these forms of violence. Namely, the current content of the item sets implies that only certain underlying ranges of these forms of IPV are observable. As a result, estimated trends in these forms of IPV—even if they are shown to be measurement invariant—may inaccurately capture true underlying trends. For example, if reductions in sexual IPV using measured physical tactics occurs alongside increases in sexual IPV using unmeasured non-physical tactics, observed rates of sexual IPV will appear to decline, when, trends in the totality of sexual IPV are stable or increasing. Indeed, focused studies using more comprehensive measures confirm the high levels of forms of sexual IPV (26) and psychological IPV (27) that the DHS does not measure. Hence, expanding these item sets is needed to capture the full range of relevant behaviors for accurate monitoring of SDG5.2.1. Such an effort need not result in large item sets, because the process of psychometric assessment can identify a precise subset that is reasonably content valid. Therefore, we recommend desk reviews of validated instruments and qualitative research in diverse settings to generate expanded item pools for sexual and psychological IPV, cognitive testing of these expanded item pools, repeated cross-cultural pilot surveys, and rigorous psychometric assessment to identify item sets that are content valid and measurement invariant across-context and across-time. Such an effort would round out the much-needed evidence to identify a common, validated item pool for inclusion in national surveys of violence against women. Agencies like the United Nations (UN), national governments, and global donors would have the evidence needed to make maximally informed decisions about the allocation of resources to prevent and to respond to IPV, based on trends in all domains of IPV that are optimally measured.

## Conclusion

This analysis is the most comprehensive assessment of the global cross-country cross-time invariance of seven physical-IPV items and five controlling-behaviors items. While measures of controlling behaviors, psychological IPV, and sexual IPV are improved, the physical IPV items are reasonable for monitoring trends in IPV against women to guide resources for effective prevention and response.

## Data Availability

Data from the Demographic and Health Surveys (DHS) are publicly available upon reasonable request to Measure DHS: https://dhsprogram.com/data/new-user-registration.cfm. Investigators must request from Measure DHS access to the data used in this analysis.

## Supporting Information

**S1 Table**. Item sets capturing physical, sexual, and psychological intimate partner violence from the Domestic Violence Module for the Demographic Health Survey Versions 5 to 7.

**S2 Table**. Invariant thresholds and loadings from alignment optimization analysis of physical intimate partner violence items and controlling behaviors items using the full and subsetted pooled samples of Demographic Health Surveys.

## References

1. World Health Organization on behalf of the United Nations Inter-Agency Working Group on Violence Against Women Estimation and Data (UNICEF U, UNODC, UNSD, UNWomen). Violence against women prevalence estimates 2018: Global, regional and national prevalence estimates for intimate partner violence against women and global and regional prevalence estimates for non-partner sexual violence against women. Geneva: World Health Organization; 2021.

2. Grose RG, Roof KA, Semenza DC, Leroux X, Yount KM. Mental health, empowerment, and violence against young women in lower-income countries: A review of reviews. Aggression and Violent Behavior 2019;46:25–36.

3. Stubbs A, Szoeke C. The effect of intimate partner violence on the physical health and health-related behaviors of women: A systematic review of the literature. Trauma, Violence, & Abuse 2021:1524838020985541.

4. Klencakova LE, Pentaraki M, McManus C. The Impact of Intimate Partner Violence on Young Women’s Educational Well-Being: A Systematic Review of Literature. Trauma, Violence, & Abuse 2021:15248380211052244.

5. Yount KM, DiGirolamo AM, Ramakrishnan U. Impacts of domestic violence on child growth and nutrition: A conceptual review of the pathways of influence. Social Science & Medicine 2011;72:1534–54.

6. Widom CS, Wilson HW. Intergenerational transmission of violence. Violence and mental health 2015:27–45.

7. Hoeffler A. What are the costs of violence? Politics, Philosophy & Economics 2017;16(4):422–45.

8. United Nations. The Sustainable Development Goals Report 2016. New York: United Nations; 2016.

9. Martín-Fernández M, Gracia E, Lila M. Psychological intimate partner violence against women in the European Union: a cross-national invariance study. BMC Public Health 2019;2019(1):1–11.

10. Martín-Fernández M, Gracia E, Lila M. Ensuring the comparability of cross-national survey data on intimate partner violence against women: a cross-sectional, population-based study in the European Union. BMJ open 2020;2020(3):e032231.

11. García-Moreno C, Jansen H, Ellsberg M, Heise L, Watts C. WHO Multi-country Study on Women’s Health and Domestic Violence against Women. Geneva, CH: World Health Organization; 2005.

12. MEASURE DHS, ICF International. Domestic Violence Module: Demographic and Health Surveys Methodology. Calverton, MD: MEASURE DHS/ICF International; 2014.

13. Yount KM, Cheong YF, Khan Z, Bergenfeld I, Kaslow N, Clark CJ, et al. Global Measurement of Physical Intimate Partner Violence to Monitor Sustainable Development Goal 5. medRxiv. 2021:2021.07. 01.21259594.

14. Basile KC, Smith SG, Breiding MJ, Black MC, Mahendra RR. Sexual Violence Surveillance: Uniform Definitions and Recommended Data Elements, Version 2.0.. Atlanta, GA: National Center for Injury Prevention and Control, Centers for Disease Control and Prevention; 2014.

15. Breiding MJ, Basile KC, Smith SG, Black MC, Mahendra RR. Intimate partner violence surveillance: uniform definitions and recommended data elements, version 2.0.. Atlanta, GA: US Department of Health and Human Services, CDC. National Center for Injury Prevention and Control; 2015.

16. Marsh HW, Hau K-T, Balla JR, Grayson D. Is more ever too much? The number of indicators per factor in confirmatory factor analysis. Multivariate behavioral research 1998;1998(2):181–220.

17. Bandalos DL, Finney SJ. Factor analysis: Exploratory and confirmatory. The reviewer’s guide to quantitative methods in the social sciences. New York: Routledge, 2018. 98–122.

18. Satorra A, Bentler PM. Ensuring positiveness of the scaled difference chi-square test statistic. Psychometrika 2010;2010(2):243–8.

19. Satorra A. Scaled and adjusted restricted tests in multi-sample analysis of moment structures. Innovations in multivariate statistical analysis: Springer; 2000. p. 233–47.

20. StataCorp. Stata Statistical Software: Release 17. College Station, TX: StataCorp LLC; 2021.

21. Muthén LK, Muthén BO. Mplus User’s Guide. Eighth Edition. Los Angeles, CA: Muthén & Muthén; 1998-2017.

22. Yount KM, Cheong YF, Khan Z, Bergenfeld I, Kaslow N, Clark CJ. Global Measurement of Partner Violence to Monitor Sustainable Development Goal 5. BMC Public Health. accepted.

23. Bishop C, Bettinson V. Evidencing domestic violence*, including behaviour that falls under the new offence of ‘controlling or coercive behaviour’. The International Journal of Evidence & Proof 2018;2018(1):3–29.

24. Myhill A. Measuring domestic violence: Context is everything. Journal of gender-based violence 2017;2017(1):33–44.

25. Basile KC, Smith SG, Breiding MJ, Black MC, Mahendra R. Sexual Violence Surveillance: Uniform Definitions and Recommended Data Elements. Atlanta, GA: National Center for Injury Prevention and Control, Centers for Disease Control and Prevention; 2014.

26. Yount K, Cheong YF, Bergenfeld I, Minh TH, Trang QT, Sales J. Impact of a Sexual Violence Prevention Edutainment Program on Sexually Violent Behavior: Randomized Controlled Trial of Global Consent Among University Men in Vietnam. nd.

27. Yount KM, Cheong YF, Miedema S, Naved RT. Development and validation of the economic coercion scale 36 (ECS-36) in rural Bangladesh. Journal of Interpersonal Violence 2021:0886260520987812.

